# Persisting inequalities in past-year drug use by sexual identity: trends across England’s 2007, 2014 and 2024 Adult Psychiatric Morbidity Surveys

**DOI:** 10.64898/2026.11.05.26367385

**Authors:** Yuting Wu, Jayati Das-Munshi, John Strang, Emmert Roberts, Amal Khanolkar, Laia Becares, Sally McManus

**Affiliations:** Department of Psychological Medicine, Institute of Psychiatry, Psychology & Neuroscience, King’s College London, London, UK; Population Health Improvement (PHI-UK), UK; National Addiction Centre, King’s College London, London, UK; South London and the Maudsley NHS Foundation Trust, London, UK; Department of Primary Care & Data Science, School of Lifecourse & Population Health Sciences, King’s College London, London, UK; Department of Global Health & Social Medicine, School of Global Affairs, King’s College London, London, UK; School of Health and Medical Sciences, City St George’s, University of London, London, UK; National Centre for Social Research, London, UK

**Keywords:** Sexual identity, LGBQ+, Drug use, Health inequalities, Population survey, England

## Abstract

**Introduction:** Sexual minority people experience disproportionate discrimination and minority stress, contributing to poorer health and health-related behaviours, including drug use. Increasing drug potency and adulteration may heighten associated harms. UK evidence on sexual identity inequalities in drug use is dated, and whether inequalities have changed over time is unclear.

**Methods:** Adult Psychiatric Morbidity Surveys 2007 (N=7403), 2014 (N=7546) and 2024 (N=6912) provide repeated cross-sectional probability samples of the household population of England. Weighted prevalence of past-year drug use was analysed by sexual identity, sex and age. Logistic regression adjusted for age, sex and area deprivation. Trends focused on 16-64-year-olds.

**Results:** In 2024, past-year drug use among adults 16+ was reported by: 9.1% heterosexual, 18.3% gay/lesbian, 26.8% bisexual, 28.3% other identity adults. Among 16-64-year-olds, drug use in sexual minority adults (2007=22.9%, 2014=27.5%, 2024=24.9%) remained at least double that of heterosexual adults (2007=10.3%, 2014=9.9%, 2024=11.6%), with adjusted odds ratios of 2.4 (95% CI:1.7-3.2, 2007), 3.2 (2.2-4.8, 2014) and 2.2 (1.4-3.4, 2024). The gap between sexual minority and heterosexual women appeared to widen, although confidence intervals were wide.

**Conclusions:** Sexual identity inequalities in drug use persisted over 17 years in England. Prevention, treatment and policies should address these inequalities and their social determinants including discrimination and exclusion. Routine drug statistics should be reported by sexual identity. Health surveys should collect gender identity and be large enough to enable disaggregation of sexual and gender identities.

**Highlights:**

- Past-year drug-use was more common in sexual minority than heterosexual adults
- Overall, odds remained at least double in sexual minority adults over the 17 years
- Sexual identity inequities evident for a range of drug types
- Among women, sexual identity inequities in drug use may have widened
- Health survey samples should be sufficient to explore age and sex intersections

## 1. Introduction

Sexual minority people experience persistent inequalities in health and health-related behaviours compared with heterosexual people (King et al., 2008; Plöderl & Tremblay, 2015). They are also disproportionately exposed to experiences of discrimination, harassment and victimisation related to their sexual identity (Katz-Wise & Hyde, 2012). The minority stress framework proposes that these experiences, alongside expectations of rejection, concealment of sexual identity and internalised stigma, can generate additional and chronic stress among sexual minority people (Meyer, 2003). Such stressors may contribute to poorer social and health outcomes, and drug use has been proposed as one potential coping response (Parent et al., 2019). Drug use, in this analysis, refers to the use of a substance for a purpose not consistent with legal or medical guidelines, for example the non-medical use of prescription medications or the recreational use of illicit drugs. Such use of drugs is associated with a wide range of health and social harms, including premature mortality. In 2024, 5,565 deaths related to drug poisoning were registered in England and Wales, the highest number recorded since records began in 1993 (ONS 2025). These risks may be heightened by increases in the potency of some drugs. Cocaine sold to users in Europe in 2024 was 44% purer than in 2014 (EUDA, 2026), while high-strength cannabis has increasingly displaced weaker varieties in Britain (Freeman et al., 2021; Potter et al., 2018). Nitazenes, synthetic opioids generally far more potent than heroin, have entered the heroin and counterfeit medicine markets (Holland et al., 2024), with implications for drug-related mortality (Chen et al., 2026).

Drug use is consistently reported to be more prevalent among sexual minority than heterosexual people. A recent meta-analysis of 304 studies found that sexual minority youth had consistently higher odds of using a range of illicit drugs than their heterosexual peers. Across substances, odds ranged from 71% higher for cannabis use (OR 1.71) to 363% higher for heroin use (OR 4.63), with wider inequities among plurisexual youth, sexual minority girls and young women, and those aged 18 or under (Mereish et al., 2026). Recent evidence from the UK Millennium Cohort Study similarly found higher odds of drug and cannabis use among sexual minority adolescents, with inequalities varying by ethnicity (Khanolkar et al., 2023). Among adults, analyses of national probability samples find similar patterns. In the National Survey on Drug Use and Health (NSDUH), substantially higher past-year illicit drug use was reported by gay, lesbian and especially bisexual adults in the United States (SAMHSA, 2016, 2023). Crime Survey for England and Wales (CSEW) data for 2011/12-2013/14 found past-year drug use of 28.4% among lesbian, gay and bisexual adults compared with 8.1% among heterosexual adults aged 16-59. This difference was evident for most drug types and the relative difference was greater in women (22.9% vs 5.1%) than men (33.0% vs 11.1%) (Home Office, 2014). Analyses of the combined 2007 and 2014 Adult Psychiatric Morbidity Survey (APMS) samples found higher rates of self-reported past year drug use among 16 to 64 year olds identifying as bisexual (37%) and lesbian or gay (25.3%) compared with those identifying as heterosexual (10.5%) or reporting another sexual identity (8.5%) (Pitman et al., 2022). These differences remained despite adjustment for reported experiences of bullying and discrimination (Pitman et al., 2022).

There are significant gaps in the evidence base. Much existing research has used convenience samples or lacked a heterosexual comparison group. Initial research on substance use among sexual minority adults often recruited via social and community groups (Hughes et al., 2020). Community-based LGBT+ surveys face related issues; the magnitude of inequities and variation by drug types cannot be reliably established from convenience data (Drabble et al., 2018). National social and health surveys have also not consistently collected information on sexual identity, and data on gender identity have been even more limited (ONS, 2019). Where sexual identity is measured in probability surveys, achieving sufficiently large samples can be challenging, given sexual minority participants tend to comprise less than 5% of general population samples (Hayes et al., 2012). Small samples also limit disaggregation by demographic and identity subgroups, despite evidence that bisexual people, and bisexual women in particular, may experience particularly pronounced inequalities (Schuler & Collins, 2020).

Another limitation is that much of the evidence base is dated and does not tell us about change over time or differences by drug type. The most recent CSEW reporting of drug use by sexual identity was in 2011-2014 (ONS, 2024), and previous APMS analyses covered up to 2014. It is not clear whether sexual identity inequities observed a decade ago have persisted, narrowed, or widened over time. This has been a period marked by substantial social and legal change, alongside shifts in drug markets and the COVID-19 pandemic. Public acceptance of same-sex relationships has increased substantially over recent decades (NatCen, 2023). Legislative changes have included the introduction of civil partnerships under the Civil Partnership Act 2004, protections against discrimination on the basis of sexual orientation under the Equality Act 2010, and the legalisation of same-sex marriage in England and Wales under the Marriage (Same Sex Couples) Act 2013. Such changes may have altered exposure to discrimination-related stressors and patterns of drug use, potentially affecting the magnitude of sexual identity inequalities over time.

The limited trend evidence available is inconsistent. Analyses of the US National Survey of Family Growth (2002-2015) suggest narrowing sexual-orientation inequities in substance use among males but persistence or widening among females (Paschen-Wolff et al., 2019). Previous APMS analyses found no significant interaction between 2007 and 2014 (Pitman et al., 2022). Comparable evidence from the UK and Europe remains limited. A UK analysis covering 2014-2021 found little evidence of sustained widening, but only examined nicotine vaping, tobacco smoking and alcohol rather than illicit drug use (Bai et al., 2026).

Official UK estimates of drug use tend to be drawn from the CSEW, a survey of crime rather than of health. The CSEW is a victimisation survey administered alongside questions on being the victim of crime. This framing may position drug use as offending behaviour, potentially reducing the likelihood of disclosure. Its drug use module has historically been restricted to respondents aged 16-59 (ONS, 2024). A recent review of UK secondary data sources concluded that no existing health survey currently supports robust analysis of sexual identity differences in drug use (Davies et al., 2022).

Most of the existing literature on drug use and sexual identity, including the main meta-analytic synthesis, focuses on young people (Mereish et al., 2026) or school and college samples (Mereish, 2019; Grigsby et al., 2026). Despite evidence that inequities vary across the life course and may follow different age patterns for men and women (Schuler et al., 2018; Parent et al., 2019), substance use among sexual minority adults in mid and later life has rarely been examined in population-based samples. Because APMS 2014 only asked sexual identity of participants aged 16 to 64, Pitman and colleagues pooled analysis focused on this age group. Across the full adult age range, population-based evidence on sexual identity inequities in drug use is lacking.

The present study aims to address these limitations. We analysed the APMS, a probability sample health survey of the household population of England aged 16 and over, in which sexual identity and illicit drug use were measured in a comparable way at three points in time: 2007, 2014 and 2024 (Morris et al., 2025). We examined self-reported past-year illicit drug use among sexual minority and heterosexual participants, addressing whether drug use inequities changed over time and if trends differed by sex and age.

Specifically, we asked: (i) how large were sexual identity inequalities in past-year drug use in England in 2024, overall and by drug type; (ii) have inequalities narrowed, persisted or widened between 2007, 2014 and 2024; and (iii) whether trends differ by sex and age group.

## 2. Methods

### 2.1 Study design and participants

We conducted a secondary analysis of the cross-sectional 2007, 2014 and 2024 APMS, which covered the household population of England aged 16 years and older. Sampling used a stratified, multistage random sampling design, based on the Small User Postcode Address File. This involved the selection of primary sampling units, addresses within selected primary sampling units, and one adult from each address.

Participants had to be able to speak English sufficiently well to be interviewed, and provided verbal informed consent. Interviews were conducted in participants’ homes (or elsewhere, if preferred) by trained research interviewers, and averaged 1.5 hours. Interviews comprised computer-assisted personal interviewing, with some sensitive information collected through computer-assisted self-completion interview in which participants used the interviewer’s laptop. Further methodological detail is published elsewhere (McManus et al., 2020; Morris et al., 2025).

### 2.2 Measures

Drug use: questions were asked in the self-completion part of the interview (Roberts et al., 2016). They covered lifetime and past-year use of over twenty types of named drugs. Alongside the main drug name, a variety of informal terms were provided to help with recognition, for example ‘(marijuana, grass, hash, ganja, draw, skunk, weed, spliff)’ was listed in brackets after cannabis. The drugs asked about largely corresponded to those included in recent waves of the CSEW, with some additional drug types such as volatile substances (described on the survey as ‘glues, solvents, gas or aerosols (to sniff)’) and amyl nitrite. New psychoactive substances (NPS) as a distinct category was not included, although some of the drugs asked about (such as mephedrone), could historically be classified as NPS. Two combined drug use in the past year variables were derived: one comprised drug types asked about in every survey, including cannabis, amphetamines, cocaine powder, crack cocaine, ecstasy, heroin, methadone, tranquillisers, alkyl nitrites, anabolic steroids, volatile substances, LSD and magic mushrooms. The other captured the expanded list of drug types asked about in 2024, which additionally included nitrous oxide, methamphetamine, synthetic cannabinoids, khat and prescription opioids. In addition, trends in past-year use of seven specific drug types - cannabis, cocaine, ketamine, ecstasy/MDMA, nitrites/poppers, mephedrone, and methamphetamine - are also presented.

Sexual identity: was derived using an approach consistent with that of Pitman and colleagues (2022). In 2007 participants were randomised (50:50) to one of two versions of a question, derived from American (Remafedi et al., 1992) or New Zealand (Fergusson et al., 2005) instruments. The first version asked participants ‘*Which statement best describes your current sexual orientation? This means sexual feelings, whether or not you have had any sexual partners*.’ It presented the following six options: entirely heterosexual (attracted to persons of the opposite of sex); mostly heterosexual, some homosexual feelings; bisexual (equally attracted to men and women); mostly homosexual, some heterosexual feelings; entirely homosexual (attracted to persons of the same sex); other sexual identities. The second version asked participants *‘Please choose the answer below that best describes how you think of yourself.’* It presented these six options: completely heterosexual; mainly heterosexual; bisexual; mainly gay or lesbian; completely gay or lesbian; other sexual identities. The question using ‘gay or lesbian’ elicited higher reporting of minority orientation than the question using ‘homosexual’, confirming that reporting is sensitive to question wording (Hayes et al., 2012). The wording in 2014 and 2024, however, aligned with the Office for National Statistics (ONS) national harmonised standard: heterosexual or straight; gay or lesbian; bisexual; other sexual identities (ONS, 2019). Sexual identity was asked of all participants (aged ≥16 years) in the 2007 and 2024 surveys, but only of participants aged 16–64 years in the 2014 survey. Data were collected using computer-assisted self-interviewing (CASI).

To derive broadly comparable groups, we first combined data from 2007 across the two question versions to form six categories: heterosexual; mainly heterosexual; bisexual; mainly homosexual; homosexual; other sexual identities. Following Pitman and colleagues (2022), we then combined ‘mainly heterosexual’ with ‘heterosexual’ and ‘mainly homosexual’ with ‘lesbian/gay’. This mapped the additional ‘mainly’ categories available in 2007 onto the closest corresponding categories available in later surveys and avoided disproportionately enlarging the sexual minority group by including the relatively large ‘mainly heterosexual’ category. While there are differences in practice, this approach was used to approximate comparable groupings over time. Due to the small numbers in some categories, a binary sexual identity variable (heterosexual vs sexual minority) was also generated.

Sociodemographic factors: These included age (16-24, 25-34, 35-44, 45-54, 55-64, 65+ years), sex (male, female), and area-level deprivation, measured using Index of Multiple Deprivation (IMD) quintiles assigned according to participants’ residential postcode. Although gender identity was asked on the most recent (2024) survey, a binary coded men/women was the only classification available across all the surveys.

### 2.3 Statistical analysis

Analyses were weighted and accounted for complex design, selection probabilities (likelihood that someone would be selected to take part), and non-response. Population control totals drew on the UK Office for National Statistics (ONS) population estimates for age by sex and region. Analyses were conducted in Stata 19 MP. Participants with missing data on sexual identity or drug use (topics covered in the self-completion section of the interview) were excluded from the relevant analyses; unweighted bases in each table therefore vary and reflect complete-case analysis for each measure.

Using the 2024 survey, we described the distribution of sexual identity across four categories and the prevalence of any past-year illicit drug use within each category. The distributions of any drug use and seven specific drug types across survey years were also examined. We then assessed patterns and trends in past-year drug use by binary sexual identity, for both any drug use and the seven specific drug types, with estimates additionally stratified by sex and age group.

For all descriptive analyses, unweighted bases and numbers of cases were reported alongside weighted prevalence estimates with 95% confidence intervals (CIs), where applicable. Analyses involving sexual identity across all three survey years were restricted to participants aged 16-64 years because sexual identity was assessed only in this age range in APMS 2014. Such analyses across the full adult age range (including those aged ≥65 years) were therefore available only for 2007 and 2024.

Logistic regression was used to examine the association between sexual identity and any past-year drug use. Models were fitted separately for participants aged 16-64 years in 2007, 2014 and 2024, with both unadjusted models and models adjusted for age, sex and IMD reported.

## 3. Results

### 3.1 Past-year drug use by sexual identity in 2024

The APMS samples comprised 7403 participants in 2007, 7546 in 2014, and 6912 in 2024.

In 2024, 81.9% of participants identified as heterosexual or straight, around 5% identified as bisexual, gay or lesbian, or other sexual identities (3.1%, 1.9%, and 0.4%, respectively); and sexual identity was missing or unavailable for 12.8% (Supplementary Table S1). In 2024, one in ten (10.2%; 95% CI 9.0, 11.4) adults aged 16 and over reported illicit drug use in the past year, on a measure comparable across survey years (Supplementary Table S2). Past-year illicit drug use was lowest among heterosexual adults (9.1%; 95% CI 7.9, 10.4), around double among gay or lesbian adults (18.3%; 6.6, 30.0), and almost triple among bisexual adults (26.8%; 17.0, 36.5) and those reporting other sexual identities (28.3%; 3.3, 53.3) (Figure 1 and Supplementary Table S1). Note that few participants reported other sexual identities (N=20), there are wide confidence intervals and the results for this group should be interpreted with caution. Estimates on an expanded measure that included drug types added to the survey in 2024 were slightly higher, but the difference between the groups was similar. Results using the binary sexual identity measure were consistent (Supplementary Table S3).

**Fig. 1.**
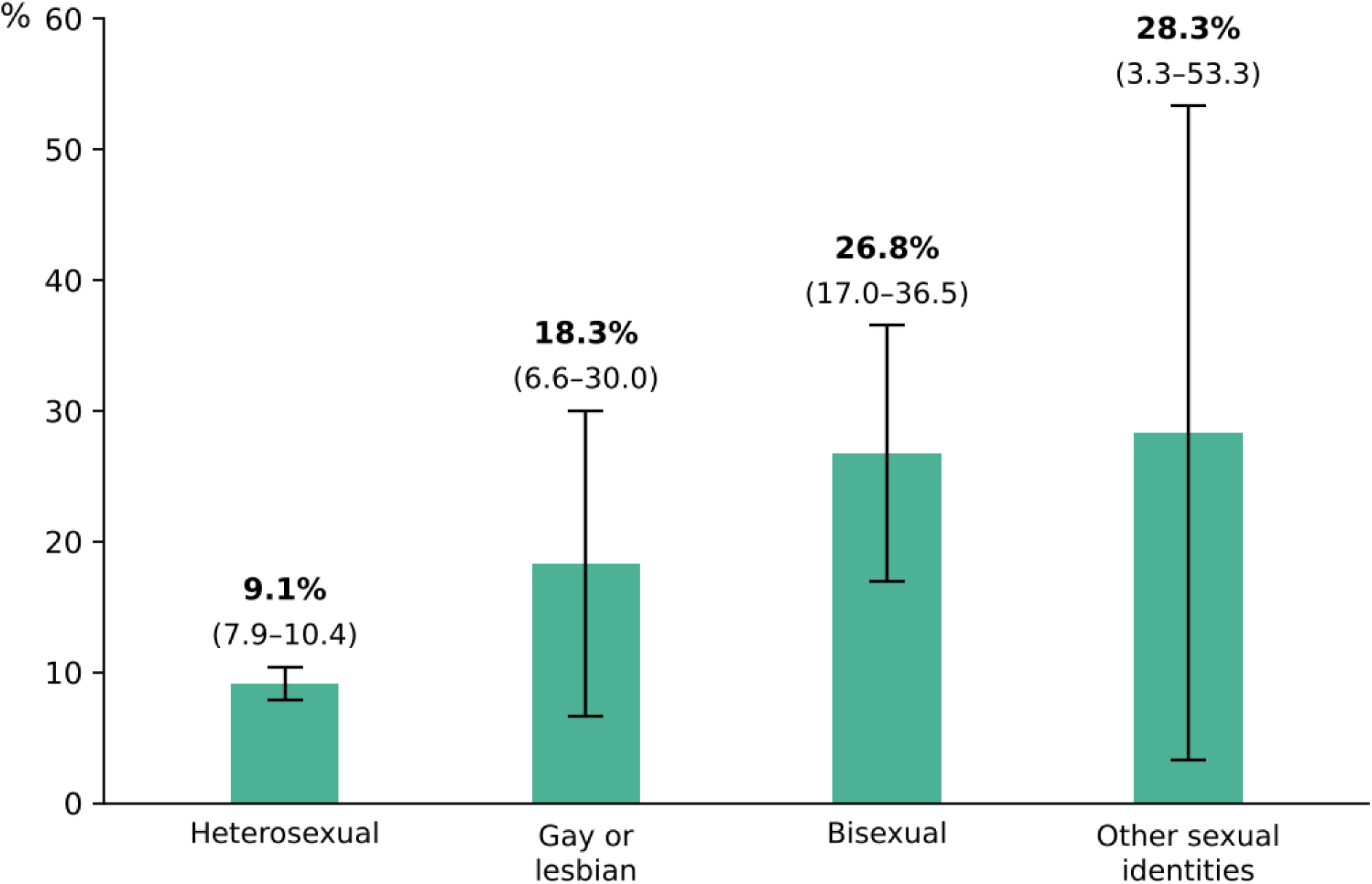
Prevalence of past-year drug use by sexual identity among adults aged 16+, APMS 2024

Cannabis was the most commonly used drug among both heterosexual (7.8%; 6.7, 9.0) and sexual minority adults (21.1%; 14.5, 27.7) (Supplementary Table S3). The prevalence of use was higher among sexual minority than heterosexual adults for all individual drug types examined, with particularly pronounced inequities for some drugs. Prevalence was more than twice as high for cocaine (5.3% vs 2.1%), compared with around five times as high for ecstasy/MDMA (5.8% vs 1.1%) and four to five times as high for ketamine (3.6% vs 0.8%). The inequity was especially large for nitrites/poppers (3.6% vs 0.1%). For these less prevalent drug types, the number of sexual minority participants reporting use was small and confidence intervals were wide.

### 3.2 Trends in past-year drug use by sexual identity

Trends across the three surveys can be examined for adults aged 16 to 64 (Table 1 and Figure 2). In this age group, past-year drug use among heterosexual adults changed little over the period (10.3% in 2007, 9.9% in 2014, 11.6% in 2024). Among sexual minority adults it was also broadly stable: 22.9% (18.3, 27.5) in 2007, 27.5% (20.2, 34.8) in 2014, and 24.9% (18.1, 31.8) in 2024. At each time point, the rate in the sexual minority group was at least twice that in the heterosexual group, alongside non-overlapping confidence intervals there was no indication that this inequity narrowed or widened over time.

**Fig. 2.**
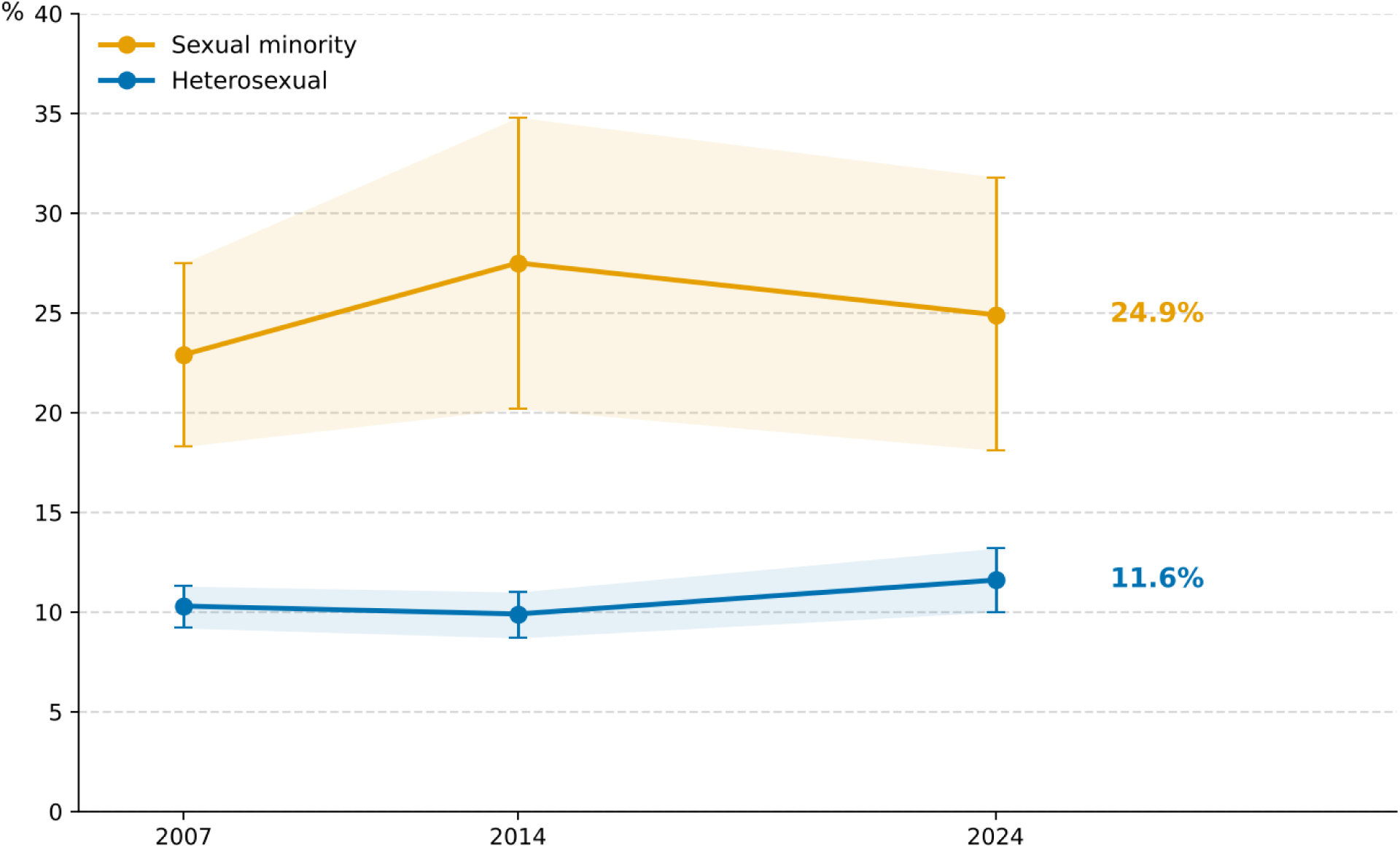
Trends in prevalence of past-year drug use by sexual identity among adults aged 16-64, APMS 2007, 2014, 2024

**Table 1.** Drug use in the past year among adults aged 16 to 64 by sexual identity, APMS 2007, 2014 and 2024.

| **Sexual identity** | **2007**<br>% (95% CI) n/N | **2014**<br>% (95% CI) n/N | **2024**<br>% (95% CI) n/N |
|---|---|---|---|
| **Any illicit drug use (comparable with 2007)** |  |  |  |
| Heterosexual | 10.3 (9.2, 11.3) 430/4,970 | 9.9 (8.7, 11.0) 388/4,692 | 11.6 (10.0, 13.2) 374/3,623 |
| Sexual minority | 22.9 (18.3, 27.5) 85/411 | 27.5 (20.2, 34.8) 58/220 | 24.9 (18.1, 31.8) 51/246 |
| **Any illicit drug use (2024 expanded)** |  |  |  |
| Heterosexual | .. | .. | 12.0 (10.4, 13.6) 391/3,623 |
| Sexual minority | .. | .. | 25.2 (18.3, 32.1) 53/246 |
| **Cannabis** |  |  |  |
| Heterosexual | 8.4 (7.4, 9.5) 341/4,971 | 8.2 (7.1, 9.2) 326/4,834 | 9.9 (8.4, 11.4) 314/3,627 |
| Sexual minority | 18.0 (13.6, 22.5) 66/411 | 21.2 (14.4, 27.9) 43/222 | 22.0 (15.1, 28.8) 42/246 |
| **Cocaine** |  |  |  |
| Heterosexual | 2.7 (2.2, 3.3) 111/4,971 | 2.2 (1.6, 2.8) 87/4,834 | 2.7 (1.9, 3.5) 85/3,627 |
| Sexual minority | 7.1 (3.8, 10.3) 23/411 | 6.2 (1.6, 10.9) 12/222 | 5.6 (1.6, 9.5) 11/246 |
| **Ketamine** |  |  |  |
| Heterosexual | .. | 0.5 (0.2, 0.9) 14/4,834 | 1.1 (0.5, 1.7) 22/3,627 |
| Sexual minority | .. | 2.0 (0.0, 5.2) 2/223 | 3.8 (0.0, 7.6) 6/246 |
| **Ecstasy/MDMA** |  |  |  |
| Heterosexual | 1.3 (1.0, 1.6) 53/4,971 | 1.8 (1.2, 2.3) 58/4,834 | 1.5 (0.8, 2.2) 37/3,627 |
| Sexual minority | 3.6 (1.4, 5.7) 14/411 | 4.9 (0.5, 9.3) 9/222 | 6.2 (1.4, 10.9) 12/246 |
| **Nitrites/poppers** |  |  |  |
| Heterosexual | 0.7 (0.4, 0.9) 29/4,971 | 0.4 (0.1, 0.6) 10/4,834 | 0.1 (0.0, 0.2) 5/3,627 |
| Sexual minority | 3.4 (1.5, 5.4) 14/411 | 5.7 (1.2, 10.2) 12/223 | 3.6 (0.1, 7.0) 6/246 |
| **Mephedrone** |  |  |  |
| Heterosexual | .. | 0.3 (0.1, 0.5) 12/4,834 | 0.0 (0.0, 0.1) 1/3,627 |
| Sexual minority | .. | 4.4 (0.1, 8.8) 7/223 | 0.5 (0.0, 1.4) 1/246 |
| **Methamphetamine** |  |  |  |
| Heterosexual | .. | .. | 0.0 0/3,627 |
| Sexual minority | .. | .. | 0.5 (0.0, 1.4) 1/246 |

‘..’ = not asked.

Trends by drug type were also broadly similar in the two groups (Table 1). Around one in five sexual minority adults used cannabis at each wave (18.0%, 21.2%, 22.0% in 2007, 2014 and 2024, respectively), as did around one in twelve heterosexual adults (8.4%, 8.2%, 9.9%), and cocaine use changed little in either group. Two exceptions were evident. First, use of nitrites (poppers) declined among heterosexual adults, from 0.7% (0.4, 0.9) in 2007 to 0.1% (0.0, 0.2) in 2024, while remaining between 3% and 6% among sexual minority adults, widening the relative difference between the groups. Second, ketamine use (first asked about in 2014) increased in both groups: from 0.5% to 1.1% among heterosexual adults, and from 2.0% to 3.8% among sexual minority adults, although the numbers of sexual minority participants reporting ketamine use were small and the confidence intervals wide.

### 3.3 Association between sexual identity and past-year drug use

In unadjusted models, sexual minority adults had higher odds of past-year illicit drug use than heterosexual adults in each survey year: 2007 (OR 2.6; 95% CI 2.0–3.5), 2014 (OR 3.5; 95% CI 2.4–5.1), and 2024 (OR 2.5; 95% CI 1.7–3.8) (Supplementary Table S4), consistent with the patterns observed in the descriptive analyses. These associations remained after adjustment for age, sex and area-level deprivation: 2007 (OR 2.4; 95% CI 1.7-3.2), 2014 (OR 3.2; 2.2-4.8), and 2024 (OR 2.2; 1.4-3.4) (Figure 3; Supplementary Table S4).

**Fig. 3.**
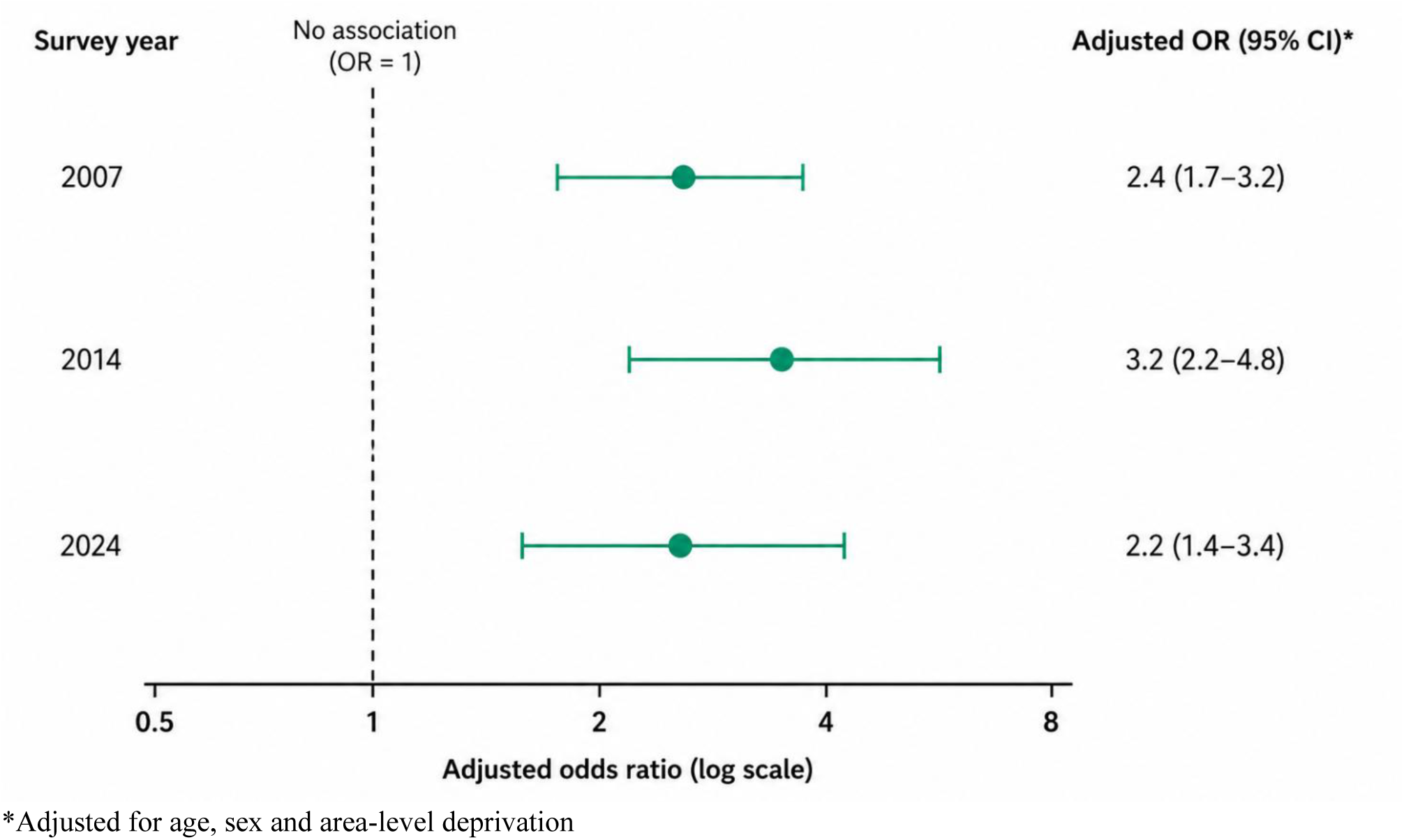
Adjusted logistic regression of the association between sexual identity and past-year drug use among adults aged 16-64, APMS 2007, 2014, 2024

### 3.4 Trends in past-year drug use by sexual identity, stratified by sex

Among adults aged 16 and over, men were nearly twice as likely as women to have used drugs in the past year across all three surveys, with rates stable over time in both sexes (12.0%, 11.3% and 13.4% for men; 6.7%, 6.0% and 7.0% for women in 2007, 2014 and 2024, respectively) (Supplementary Table S5).

Trends by sexual identity, however, appeared to move in different directions for men and women (Figure 4; Table 2). Among heterosexual men aged 16 and over, past-year drug use was 11.2% (9.7, 12.7) in 2007 and 13.2% (11.0, 15.3) in 2024. Among sexual minority men, the point estimate fell from 23.1% (17.2, 29.0) in 2007 to 16.2% (7.8, 24.5) in 2024 (based on 115 participants). As a result, the difference between sexual minority and heterosexual men evident in 2007 was no longer evident in 2024, although it should be noted that the confidence interval around the 2024 estimate for sexual minority men was wide.

**Fig. 4.**
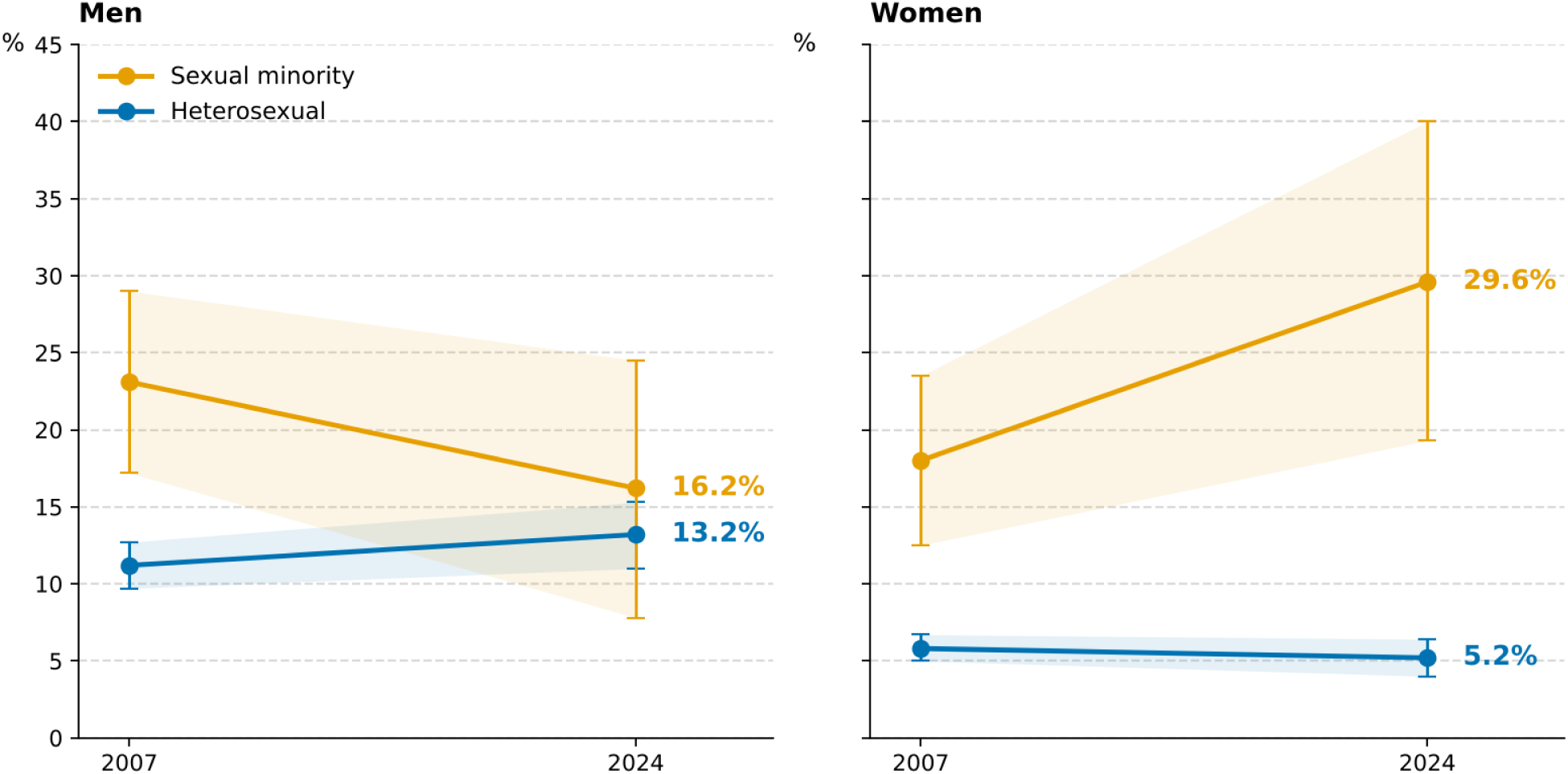
Prevalence of past-year drug use by sexual identity and sex among adults 16+, APMS 2007, 2024

**Table 2.** Drug use in the past year among men and women aged 16+ by sexual identity, APMS 2007 and 2024.

|  | **Heterosexual** |  | **Sexual minority** |  |
|---|---|---|---|---|
| **Drug use in past year** | **2007**<br>% (95% CI) n/N | **2024**<br>% (95% CI) n/N | **2007**<br>% (95% CI) n/N | **2024**<br>% (95% CI) n/N |
| **Men** |  |  |  |  |
| Any illicit drug use (comparable with 2007) | 11.2 (9.7, 12.7) 257/2,938 | 13.2 (11.0, 15.3) 236/2,390 | 23.1 (17.2, 29.0) 46/218 | 16.2 (7.8, 24.5) 18/115 |
| Any illicit drug use (2024 expanded) | .. | 13.5 (11.3, 15.7) 246/2,390 | .. | 16.4 (8.0, 24.8) 19/115 |
| Cannabis | 9.5 (8.1, 11.0) 212/2,939 | 11.2 (9.2, 13.3) 199/2,394 | 17.8 (12.2, 23.3) 35/219 | 12.6 (4.5, 20.7) 13/115 |
| Cocaine | 3.4 (2.7, 4.2) 76/2,939 | 3.4 (2.2, 4.6) 59/2,394 | 6.5 (2.2, 10.8) 10/219 | 3.4 (0.2, 6.7) 5/115 |
| Ketamine | .. | 1.2 (0.4, 2.0) 14/2,394 | .. | 0.5 (0.0, 1.3) 2/115 |
| Ecstasy/MDMA | 1.7 (1.2, 2.3) 41/2,939 | 1.9 (0.9, 2.8) 28/2,394 | 3.6 (0.4, 6.7) 7/219 | 6.3 (0.0, 13.2) 6/115 |
| Nitrites/poppers | 0.6 (0.3, 0.9) 16/2,939 | 0.1 (0.0, 0.3) 3/2,394 | 4.7 (1.6, 7.8) 11/219 | 5.1 (0.3, 9.8) 6/115 |
| Mephedrone | .. | 0.1 (0.0, 0.2) 1/2,394 | .. | 1.0 (0.0, 2.9) 1/115 |
| Methamphetamine | .. | 0.0 0/2,394 | .. | 1.0 (0.0, 2.9) 1/115 |
| **Women** |  |  |  |  |
| Any illicit drug use (comparable with 2007) | 5.8 (5.0, 6.7) 190/3,863 | 5.2 (4.0, 6.4) 163/3,217 | 18.0 (12.5, 23.5) 41/283 | 29.6 (19.3, 40.0) 34/162 |
| Any illicit drug use (2024 expanded) | .. | 5.7 (4.5, 6.9) 179/3,217 | .. | 30.0 (19.7, 40.4) 36/162 |
| Cannabis | 4.3 (3.4, 5.2) 134/3,863 | 4.5 (3.4, 5.5) 138/3,218 | 14.6 (9.2, 20.1) 33/283 | 27.4 (17.0, 37.7) 30/162 |
| Cocaine | 1.1 (0.7, 1.4) 35/3,863 | 0.8 (0.3, 1.3) 27/3,218 | 6.0 (2.1, 9.8) 13/283 | 6.9 (0.5, 13.3) 6/162 |
| Ketamine | .. | 0.5 (0.0, 0.9) 8/3,218 | .. | 6.1 (0.0, 12.7) 4/162 |
| Ecstasy/MDMA | 0.4 (0.2, 0.6) 12/3,863 | 0.4 (0.0, 0.9) 10/3,218 | 2.7 (0.6, 4.9) 7/283 | 5.5 (0.0, 11.6) 6/162 |
| Nitrites/poppers | 0.5 (0.2, 0.8) 13/3,863 | 0.1 (0.0, 0.1) 2/3,218 | 1.7 (0.0, 3.6) 4/283 | 2.4 (0.0, 7.1) 1/162 |
| Mephedrone | .. | 0.0 0/3,218 | .. | 0.0 0/162 |
| Methamphetamine | .. | 0.0 0/3,218 | .. | 0.0 0/162 |

‘..’ = not asked.

The pattern for women was the reverse. Drug use among heterosexual women was stable: 5.8% (5.0, 6.7) in 2007 and 5.2% (4.0, 6.4) in 2024, while among sexual minority women the point estimate appeared to rise from 18.0% (12.5, 23.5) in 2007 to 29.6% (19.3, 40.0) in 2024. By 2024, the rate of past-year drug use in sexual minority women was more than five times that in heterosexual women, with confidence intervals that did not overlap. This was largely accounted for by cannabis, reported by 27.4% (17.0, 37.7) of sexual minority women compared with 4.5% (3.4, 5.5) of heterosexual women; ecstasy/MDMA (5.5% vs 0.4%) and ketamine (6.1% vs 0.5%) use also contributed.

Sex-stratified unadjusted logistic regression analyses among adults aged 16–64 showed a similar pattern (Supplementary Table S4). The higher odds of past-year drug use among sexual minority men observed in 2007 (OR 2.2; 95% CI 1.5–3.2) and 2014 (OR 2.6; 95% CI 1.5–4.8) were no longer evident in 2024 (OR 1.0; 95% CI 0.5–2.0). In contrast, among women, the estimated odds ratios increased across successive surveys, from 3.5 (95% CI 2.3–5.3) in 2007 to 5.1 (3.2–8.1) in 2014 and 6.0 (3.3–10.9) in 2024.

### 3.5 Trends in past-year drug use by sexual identity, stratified by age

Drug use varied by age in every survey year, but the shape of this association has changed (Table 3). Among heterosexual adults, past-year drug use in the youngest age group (16 to 24 years) fell from 23.6% (19.5, 27.6) in 2007 to 20.6% (16.6, 24.6) in 2014 and 15.0% (9.4, 20.6) in 2024. In contrast, drug use rose in midlife: between 2007 and 2024 it increased from 6.9% (5.5, 8.4) to 12.4% (9.7, 15.1) among those aged 35 to 44, tripled among those aged 45 to 54 (from 2.9% to 8.8%), and more than doubled among those aged 55 to 64 (from 1.8% to 4.6%).

**Table 3.** Drug use in the past year by age group and sexual identity: APMS 2007, 2014, 2024.

|  | **Heterosexual** |  |  | **Sexual minority** |  |  |
|---|---|---|---|---|---|---|
| **Age group** | **2007**<br>% (95% CI) n/N | **2014**<br>% (95% CI) n/N | **2024**<br>% (95% CI) n/N | **2007**<br>% (95% CI) n/N | **2014**<br>% (95% CI) n/N | **2024**<br>% (95% CI) n/N |
| **Any drug use (comparable with 2007)** |  |  |  |  |  |  |
| 16–24 | 23.6 (19.5, 27.6) 118/498 | 20.6 (16.6, 24.6) 102/500 | 15.0 (9.4, 20.6) 34/239 | 31.2 (19.2, 43.2) 20/63 | 34.9 (18.3, 51.6) 14/38* | 34.1 (19.1, 49.2) 13/43* |
| 25–34 | 18.3 (15.4, 21.3) 158/930 | 13.8 (10.8, 16.9) 115/913 | 18.0 (13.8, 22.2) 100/650 | 33.3 (22.7, 43.8) 29/96 | 26.9 (11.3, 42.6) 13/50 | 21.8 (11.6, 31.9) 17/71 |
| 35–44 | 6.9 (5.5, 8.4) 95/1,296 | 7.8 (6.1, 9.6) 84/1,048 | 12.4 (9.7, 15.1) 106/860 | 20.5 (11.2, 29.9) 22/104 | 38.0 (21.5, 54.6) 17/44* | 12.8 (3.0, 22.5) 8/59 |
| 45–54 | 2.9 (1.9, 3.9) 35/1,037 | 4.7 (3.4, 6.1) 58/1,158 | 8.8 (6.6, 11.1) 82/837 | 10.6 (2.9, 18.3) 9/86 | 15.5 (4.4, 26.6) 8/47* | 30.6 (14.5, 46.7) 12/42* |
| 55–64 | 1.8 (1.0, 2.7) 24/1,209 | 2.7 (1.6, 3.8) 29/1,073 | 4.6 (3.2, 6.1) 52/1,037 | 8.8 (0.8, 16.8) 5/62 | 14.3 (2.2, 26.5) 6/41* | 5.9 (0.0, 17.5) 1/31* |
| 65+ | 0.9 (0.5, 1.4) 17/1,831 | .. | 1.0 (0.5, 1.5) 26/1,980 | 2.2 (0.0, 5.4) 2/90 | .. | 6.8 (0.0, 17.4) 2/32* |
| **Any drug use (2024 expanded)** |  |  |  |  |  |  |
| 16–24 | .. | .. | 15.0 (9.4, 20.6) 34/239 | .. | .. | 34.1 (19.1, 49.2) 13/43* |
| 25–34 | .. | .. | 18.7 (14.5, 22.9) 104/650 | .. | .. | 22.6 (12.3, 32.8) 18/71 |
| 35–44 | .. | .. | 12.8 (10.1, 15.5) 110/860 | .. | .. | 13.1 (3.3, 22.9) 9/59 |
| 45–54 | .. | .. | 9.3 (6.9, 11.6) 86/837 | .. | .. | 30.6 (14.5, 46.7) 12/42* |
| 55–64 | .. | .. | 5.1 (3.5, 6.6) 57/1,037 | .. | .. | 5.9 (0.0, 17.5) 1/31* |
| 65+ | .. | .. | 1.4 (0.9, 2.0) 35/1,980 | .. | .. | 8.7 (0.0, 20.0) 3/32* |

‘\*’ Note small base size for these estimates (unweighted N <50).
‘..’ = not asked.

The profile was different among sexual minority adults. Around a third of sexual minority individuals aged 16 to 24 years reported past-year drug use at every wave (31.2% in 2007, 34.9% in 2014, 34.1% in 2024), while drug use fell among young heterosexual adults (Table 3). In 2024 the rate in sexual minority 16 to 24 year olds (34.1%; 19.1, 49.2) was more than double that of their heterosexual peers (15.0%; 9.4, 20.6), whereas the two groups had been closer in 2007 (31.2% vs 23.6%). Drug use among sexual minority adults aged 45 to 54 also increased, from 10.6% (2.9, 18.3) in 2007 to 30.6% (14.5, 46.7) in 2024. However, a similar midlife rise was also observed in the heterosexual group. The divergence appears to be driven by changing use of cannabis (Table S6; Figure 5). Sexual minority estimates for individual age groups were based on fewer than 50 participants in most cells in 2014 and 2024 and should be interpreted with caution.

**Fig. 5.**
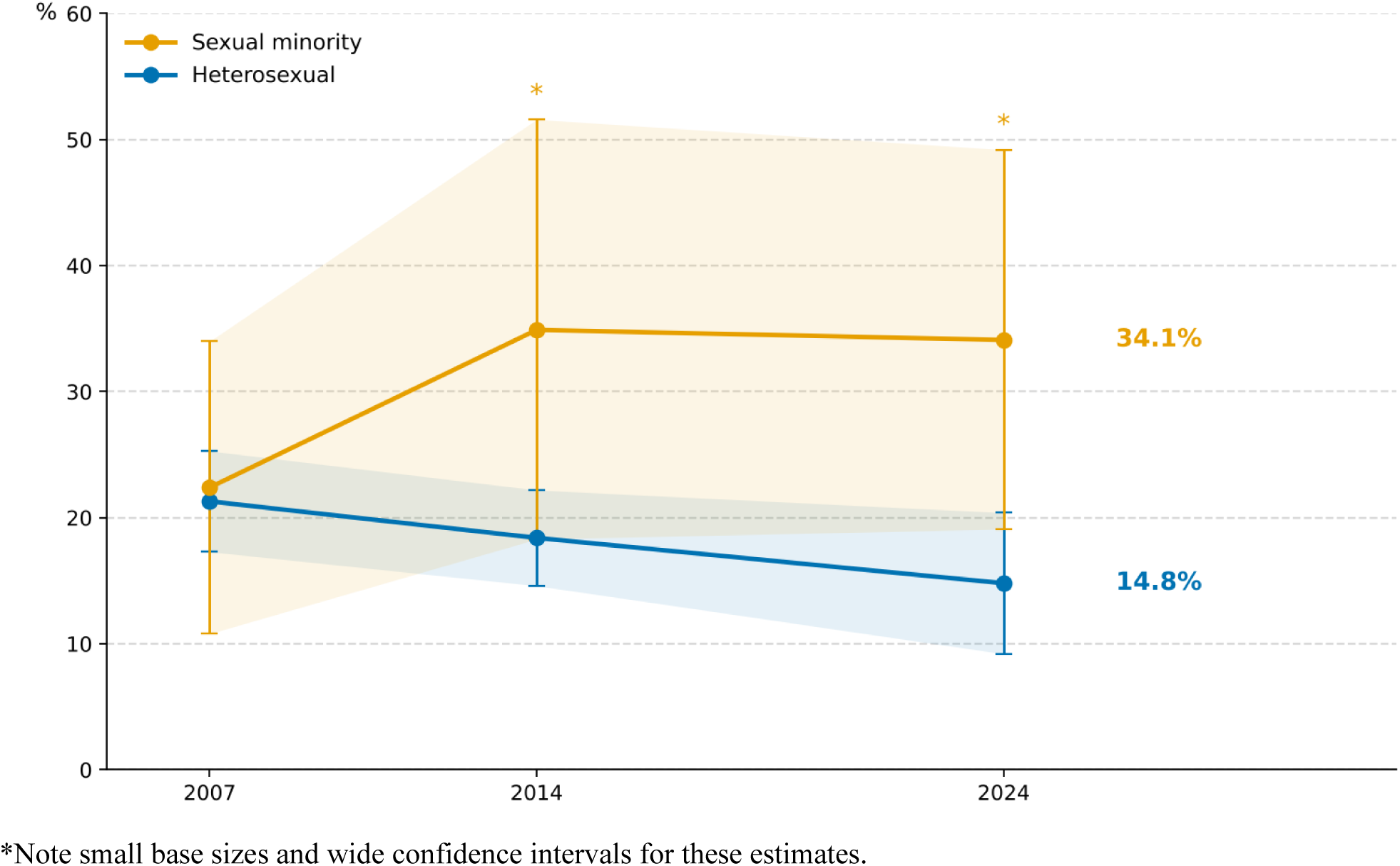
Prevalence of past-year cannabis use by sexual identity among adults aged 16-24, APMS 2007, 2014, 2024

## 4. Discussion

Past-year drug use was at least twice as common among sexual minority adults as heterosexual adults at every point (2007, 2014, 2024), with no evidence that this inequity has narrowed. In 2024, around one in four (23.9%) sexual minority adults aged 16 and over reported illicit drug use in the past year, compared with about one in eleven (9.1%) heterosexual adults, and every drug type examined was used by a higher proportion of the sexual minority group. Beneath this overall stability, trends appeared to have moved in different directions for men and women. Among men, the difference observed in 2007 was no longer apparent by 2024. Among women the pattern was reversed: drug use among sexual minority women rose from 18.0% in 2007 to 29.6% in 2024, a rate more than five times that in heterosexual women, largely accounted for by cannabis. The age profile of drug use also appears to have shifted: drug use fell among young heterosexual adults and rose in midlife in both groups, while around a third of sexual minority 16 to 24 year olds reported past-year drug use at every wave.

These findings indicate that the higher levels of drug use previously observed among sexual minority people in England have persisted through a period of substantial legislative and attitudinal change. This mirrors the persistence of inequalities across other health outcomes despite improvements in legal rights and social acceptance, which may reflect continued exposure to stigma and discrimination related to sexual identity and other minority stressors (Khanolkar, 2024). While confidence intervals were wide, the observed results suggest that drug use may be rising among sexual minority women, with no comparable increase among sexual minority men. A possible divergence in trends by sex is consistent with US data suggesting narrowing inequities among males but persistence or widening among females (Paschen-Wolff et al., 2019), and with evidence that inequalities may be particularly pronounced for bisexual women (Schuler & Collins, 2020), although the samples here were too small to disaggregate sexual identity subgroups. Sexual victimisation has been proposed as one contributing pathway, given higher reported rates among sexual minority women, particularly bisexual women (Canan et al., 2021), and evidence of greater substance use to cope with such experiences among sexual minority women (López & Yeater, 2021). Poor mental health and substance use frequently co-occur among sexual minority populations, and minority stress theory frames drug use as a means of coping with stigma, discrimination and victimisation within heteronormative social contexts (Bränström & Pachankis, 2018; Meyer, 2003). While previous APMS analyses found that these inequities were attenuated after adjustment for reported bullying and amplified after adjustment for discrimination, remaining significant in both cases (Pitman et al., 2022), social and peer norms surrounding substance use, and living in heteronormative environments may represent additional pathways contributing to these inequalities (Mereish et al., 2017). The persistence of nitrites (poppers) use among sexual minority adults, and among sexual minority men in particular, while use declined to very low levels among heterosexual adults, is consistent with evidence that poppers are particularly used by gay and bisexual men in sexual contexts (Vaccher et al., 2020; Schwartz et al., 2020). This illustrates how patterns of use can be identity specific, with implications for the substances that harm reduction services should anticipate. With drug-related deaths at record levels (ONS, 2025) and use rising in midlife, these results underline the need for drug treatment and wider health services to ask about and respond to drug use in sexual minority populations (Hillyard et al., 2026a), and among sexual minority women in particular (Hillyard et al., 2026b).

The finding that inequalities have persisted through a period in which formal legal equality was substantially extended has implications for how these inequalities are understood. If legislative change alone were sufficient, the inequities documented may be expected to have narrowed. That they have not is consistent with accounts locating drivers in everyday and structural stigma, which formal legal change addresses only indirectly. For population health policy this points away from framing sexual minority drug use as an individual behavioural risk, and towards responses that see it as a socially patterned inequality with structural determinants. The signs of diverging trends between men and women are also notable, and may reflect changing population norms around cannabis use, the growth and changing composition of the bisexual population among younger women, and different and changing exposures to minority stress.

Our study offers particular strengths over the existing evidence base. This study provides a heterosexual comparison group within a single sampling frame and mode, and spans the full adult age range from 16, including older adults who are excluded from the CSEW drug module and from many studies of sexual minority groups. This enabled analysis of trends in sexual identity inequities in drug use, and in use of specific drug types, in England over an extended period spanning major social change. Because APMS is a health survey - the government’s key official source of data on mental health in England’s adult population - drug use was measured within a health-behaviour framing, and both drug use and sexual identity were collected by computer-assisted self-completion rather than face to face, which is likely to support better disclosure of sensitive information. Trend estimates drew on a derived variable restricted to the drug types asked about in every survey, so trends are not an artefact of the expanding drug list.

However, important limitations remain. The small size of key subgroups led to imprecise estimates and prevented investigation of sexual identity subgroups, adjustment for a wider range of potential confounders, and examination of intersections between age, sex, ethnicity and sexual identity. The repeated cross-sectional design is ideal for examining change at the population level but is not suitable for causal questions. Differences were assessed by inspecting the overlap of 95% confidence intervals, a conservative approach, and formal tests of trend or of interactions (for example sex by identity by survey year) were not conducted due to the survey years being separate datasets. Drug use was self-reported, so under-reporting is possible, although the self-completion mode and health survey framing may mitigate this. The self-completion was not completed by all participants, accounting for the substantial proportion not providing a sexual identity response, increasing overall missing data and introducing potential non-response bias. Willingness to report both illicit drug use and a non-heterosexual sexual identity may also vary with their perceived social acceptability and may have changed over time (Harrison, 1995). The sexual identity question also changed between waves: participants in 2007 were randomised to one of two question versions while 2014 and 2024 used the ONS harmonised standard, and reporting of minority identity is sensitive to question wording (Hayes et al., 2012), so trends by sexual identity could partly reflect measurement change as well as changing willingness to disclose. The proportion and composition of adults identifying with a sexual minority status may itself have shifted, so within-group trends may reflect changing membership of the group as well as changing behaviour. Because sexual identity was only established for 16 to 64 year olds in 2014, all-age comparisons rest on two time points. Sex was binary coded as this is the classification available across all three surveys, and gender identity was asked only in 2024, so trans and non-binary participants could not be identified consistently. Finally, as a household survey, key groups such as people who are in prison or homeless were out of scope; these are groups likely to experience higher than average rates of drug use (Bebbington et al., 2021; Khanolkar et al., 2026).

## 5. Conclusion

Drawing on three national probability-sample health surveys spanning 17 years, this study shows that sexual minority adults in England remain more than twice as likely as heterosexual adults to have used drugs in the past year, and that this inequity is now particularly evident among sexual minority women and young people. Efforts to reduce these inequalities should consider the social and structural factors that may contribute to elevated drug use among sexual minority people, including social norms around drug use and discrimination. Drug prevention and treatment services should also be inclusive and responsive to the needs of sexual minority populations. Routine drug use statistics should collect and publish estimates by sexual identity, which the CSEW last did for 2011 to 2014, and future surveys should be designed with samples large enough to disaggregate bisexual, gay and lesbian, and other identities, and their intersections with age and sex.

## Statements and Declarations

### CRediT authorship contribution statement

Yuting Wu: Formal analysis, Investigation, Visualization, Writing - original draft. Jayati Das-Munshi: Conceptualization, Supervision, Writing - review & editing. John Strang: Conceptualization, Writing - review & editing. Emmert Roberts: Conceptualization, Writing - review & editing. Amal Khanolkar: Conceptualization, Writing - review & editing. Laia Becares: Conceptualization, Writing - review & editing. Sally McManus: Conceptualization, Methodology, Supervision, Visualization, Writing - original draft, Writing - review & editing.

### Ethical statement

APMS 2007 and APMS 2014 were approved by the Royal Free Hospital and Medical School Research Ethics Committee (06/Q0501/71) and the West London National Research Ethics Committee (14/LO/0411) respectively. Ethical approval for APMS 2023/4 was obtained from NatCen’s internal Research Ethics Committee (1 November 2022). All participants provided verbal informed consent. The present study is a secondary analysis of de-identified survey data and did not require additional ethical approval.

## Acknowledgements

We would like to thank the thousands of people across England who generously participated in the survey. NHS England commissioned APMS series with funding from the Department of Health and Social Care in England. SM acknowledges salary support from the UKRI Prevention Research Partnership (Violence, Health and Society; MR-V049879/1). This analysis was suggested by Dr Mollie Virgo, Secretariat to the Advisory Council on the Misuse of Drugs (ACMD), Home Office to inform national monitoring and strategic review.

## Data availability

The deidentified APMS datasets, data dictionary, protocol, participant materials, and full documentation are lodged with the UK Data Service archive. Permission to use the datasets was obtained from NHS England. Requests for further use should be made via the UK Data Service (https://ukdataservice.ac.uk/), and are subject to permission from NHS England.

## Declaration of Generative AI and AI-assisted technologies in the writing process

During the preparation of this work the authors used Anthropic’s Claude (Opus 4.5) and OpenAI’s ChatGPT (GPT-5.6) in order to generate plotting code from author-supplied data to produce the figures. No data were generated, imputed or altered by these tools. After using this tool/service, the authors reviewed and edited the content as needed and take full responsibility for the content of the publication.

