## Supplementary Tables S1-S6 and Figure S1 for "Persisting inequalities in past-year drug use by sexual identity: trends across England’s 2007, 2014 and 2024 Adult Psychiatric Morbidity Surveys"

### Supplementary Materials file

### Supplementary Tables

**Table S1 Sexual identity distribution among adults aged 16+, and prevalence of past-year drug use by sexual identity, APMS 2024**

| Sexual identity | Unweighted n | Weighted % (95% CI) | Any illicit drug use (comparable measure), n/N | Weighted % (95% CI) | Any illicit drug use (2024 expanded measure), n/N | Weighted % (95% CI) |
|---|---|---|---|---|---|---|
| Heterosexual | 5,617 | 81.9 (80.2–83.5) | 400/5,611 | 9.1 (7.9–10.4) | 426/5,611 | 9.5 (8.3–10.8) |
| Gay or lesbian | 100 | 1.9 (1.4–2.5) | 13/100 | 18.3 (6.6–30.0) | 13/100 | 18.3 (6.6–30.0) |
| Bisexual | 158 | 3.1 (2.5–3.8) | 34/158 | 26.8 (17.0–36.5) | 37/158 | 27.3 (17.6–37.1) |
| Other sexual identities | 20 | 0.4 (0.2–0.7) | 6/20 | 28.3 (3.3–53.3) | 6/20 | 28.3 (3.3–53.3) |
| Missing/not available* | 1,017 | 12.8 (11.4–14.3) | – | – | – | – |

*Participants who did not complete the self-completion module of the interview and therefore missing responses on both sexual identity and drug use.

**Table S2 Past-year drug use among adults aged 16+: APMS 2007, 2014, 2024**

| Past-year drug use | 2007 n/N | 2007 % (95% CI) | 2014 n/N | 2014 % (95% CI) | 2024 n/N | 2024 % (95% CI) |
|---|---|---|---|---|---|---|
| Any illicit drug use (comparable measure) | 534/7,356 | 9.2 (8.4–10.1) | 461/6,909 | 8.6 (7.6–9.5) | 487/6,135 | 10.2 (9.0–11.4) |
| Any illicit drug use (expanded 2024 measure) | .. | .. | .. | .. | 518/6,135 | 10.6 (9.4–11.8) |
| Cannabis | 414/7,394 | 7.4 (6.6–8.3) | 378/7,545 | 6.7 (5.9–7.4) | 410/6,911 | 7.9 (6.9–8.9) |
| Cocaine | 134/7,394 | 2.5 (2.0–2.9) | 100/7,545 | 1.8 (1.4–2.3) | 106/6,911 | 2.1 (1.6–2.7) |
| Ketamine | .. | .. | 16/7,546 | 0.5 (0.2–0.7) | 29/6,911 | 0.9 (0.5–1.3) |
| Ecstasy/MDMA | 67/7,394 | 1.2 (0.9–1.5) | 67/7,545 | 1.4 (1.0–1.8) | 54/6,911 | 1.3 (0.8–1.8) |
| Nitrites/poppers | 44/7,394 | 0.7 (0.5–0.9) | 22/7,546 | 0.5 (0.2–0.7) | 13/6,911 | 0.3 (0.1–0.5) |
| Mephedrone | .. | .. | 20/7,546 | 0.4 (0.2–0.6) | 2/6,911 | 0.05 (0.0–0.1) |
| Methamphetamine | .. | .. | .. | .. | 1/6,911 | 0.02 (0.0–0.1) |

‘..’ = not asked.

**Table S3 Drug use in the past year among adults aged 16+ by sexual identity, APMS 2007, 2024**

| Sexual identity | 2007<br>% (95% CI) n/N | 2024<br>% (95% CI) n/N |
|---|---|---|
| **Any illicit drug use (comparable with 2007)** | | |
| Heterosexual | 8.5 (7.6, 9.4) 447/6,801 | 9.1 (7.9, 10.4) 400/5,611 |
| Sexual minority | 20.4 (16.4, 24.5) 87/501 | 23.9 (17.3, 30.6) 53/278 |
| Total | 9.2 (8.4, 10.1) 534/7,356 | 10.2 (9.0, 11.4) 487/6,135 |
| **Any illicit drug use (2024 expanded)** | | |
| Heterosexual | .. | 9.5 (8.3, 10.8) 426/5,611 |
| Sexual minority | .. | 24.3 (17.6, 30.9) 56/278 |
| Total | .. | 10.6 (9.4, 11.8) 518/6,135 |
| **Cannabis** | | |
| Heterosexual | 6.9 (6.0, 7.7) 346/6,802 | 7.8 (6.7, 9.0) 338/5,616 |
| Sexual minority | 16.1 (12.2, 20.1) 68/502 | 21.1 (14.5, 27.7) 44/278 |
| Total | 7.4 (6.6, 8.3) 414/7,394 | 7.9 (6.9, 8.9) 410/6,911 |
| **Cocaine** | | |
| Heterosexual | 2.2 (1.8, 2.6) 111/6,802 | 2.1 (1.5, 2.7) 86/5,616 |
| Sexual minority | 6.2 (3.3, 9.1) 23/502 | 5.3 (1.5, 9.0) 11/278 |
| Total | 2.5 (2.0, 2.9) 134/7,394 | 2.1 (1.6, 2.7) 106/6,911 |
| **Ketamine** | | |
| Heterosexual | .. | 0.8 (0.4, 1.3) 22/5,616 |
| Sexual minority | .. | 3.6 (0.0, 7.2) 6/278 |
| Total | .. | 0.9 (0.5, 1.3) 29/6,911 |
| **Ecstasy/MDMA** | | |
| Heterosexual | 1.0 (0.8, 1.3) 53/6,802 | 1.1 (0.6, 1.7) 38/5,616 |
| Sexual minority | 3.1 (1.3, 5.0) 14/502 | 5.8 (1.4, 10.3) 12/278 |
| Total | 1.2 (0.9, 1.5) 67/7,394 | 1.3 (0.8, 1.8) 54/6,911 |
| **Nitrites/poppers** | | |
| Heterosexual | 0.5 (0.3, 0.7) 29/6,802 | 0.1 (0.0, 0.2) 5/5,616 |
| Sexual minority | 3.1 (1.4, 4.9) 15/502 | 3.6 (0.3, 6.9) 7/278 |
| Total | 0.7 (0.5, 0.9) 44/7,394 | 0.3 (0.1, 0.5) 13/6,911 |
| **Mephedrone** | | |
| Heterosexual | .. | 0.0 (0.0, 0.1) 1/5,616 |
| Sexual minority | .. | 0.4 (0.0, 1.3) 1/278 |
| Total | .. | 0.1 (0.0, 0.1) 2/6,911 |
| **Methamphetamine** | | |
| Heterosexual | .. | 0.0 0/5,616 |
| Sexual minority | .. | 0.4 (0.0, 1.3) 1/278 |
| Total | .. | 0.0 (0.0, 0.1) 1/6,911 |

'..' = not asked.

**Table S4 Unadjusted and adjusted logistic regression of the association between sexual identity and any past-year drug use among adults aged 16–64, APMS 2007, 2014 and 2024**

| Survey year | Analysis N | OR (95% CI) | p-value |
|---|---|---|---|
| **Model 1: Overall, unadjusted** | | | |
| 2007 | 5,381 | 2.6 (2.0–3.5) | <0.001 |
| 2014 | 4,912 | 3.5 (2.4–5.1) | <0.001 |
| 2024 | 3,869 | 2.5 (1.7–3.8) | <0.001 |
| **Model 2: Sex-stratified, unadjusted** | | | |
| Men | | | |
| 2007 | 2,345 | 2.2 (1.5–3.2) | <0.001 |
| 2014 | 1,920 | 2.6 (1.5–4.8) | 0.002 |
| 2024 | 1,555 | 1.0 (0.5–2.0) | 0.933 |
| Women | | | |
| 2007 | 3,036 | 3.5 (2.3–5.3) | <0.001 |
| 2014 | 2,992 | 5.1 (3.2–8.1) | <0.001 |
| 2024 | 2,309 | 6.0 (3.3–10.9) | <0.001 |
| **Model 3: Overall, adjusted** | | | |
| 2007 | 5,381 | 2.4 (1.7–3.2) | <0.001 |
| 2014 | 4,912 | 3.2 (2.2–4.8) | <0.001 |
| 2024 | 3,864 | 2.2 (1.4–3.4) | <0.001 |

Note: Odds ratios compare sexual minority with heterosexual adults (reference group). Model 3 was adjusted for age, sex and area-level deprivation.

**Table S5 Trends in past-year illicit drug use among adults aged 16+, by sex: APMS 2007, 2014 and 2024**

| | Male | | | Female | | |
|---|---|---|---|---|---|---|
| **Drug use in past year** | **2007** % (95% CI) n/N | **2014** % (95% CI) n/N | **2024** % (95% CI) n/N | **2007** % (95% CI) n/N | **2014** % (95% CI) n/N | **2024** % (95% CI) n/N |
| Any drug use (comparable with 2007) | 12.0 (10.5, 13.4) 303/3,174 | 11.3 (9.8, 12.9) 256/2,783 | 13.4 (11.4, 15.4) 269/2,599 | 6.7 (5.8, 7.6) 231/4,182 | 6.0 (5.1, 6.9) 205/4,126 | 7.0 (5.8, 8.2) 216/3,529 |
| Any drug use (2024 expanded) | .. | .. | 13.7 (11.7, 15.8) 280/2,599 | .. | .. | 7.5 (6.3, 8.8) 236/3,529 |
| Cannabis | 10.0 (8.6, 11.4) 247/3,192 | 8.7 (7.4, 10.0) 208/3,057 | 10.4 (8.6, 12.1) 225/2,940 | 5.0 (4.1, 5.9) 167/4,202 | 4.7 (3.9, 5.5) 170/4,488 | 5.6 (4.5, 6.6) 183/3,964 |
| Cocaine | 3.6 (2.8, 4.4) 86/3,192 | 2.6 (1.9, 3.3) 61/3,057 | 3.2 (2.2, 4.2) 69/2,940 | 1.4 (0.9, 1.9) 48/4,202 | 1.1 (0.7, 1.5) 39/4,488 | 1.1 (0.6, 1.6) 37/3,964 |
| Ketamine | .. | 0.7 (0.2, 1.2) 10/3,058 | 1.0 (0.4, 1.7) 16/2,940 | .. | 0.2 (0.0, 0.4) 6/4,488 | 0.7 (0.2, 1.3) 13/3,964 |
| Ecstasy/MDMA | 1.9 (1.3, 2.4) 48/3,192 | 1.8 (1.1, 2.5) 39/3,057 | 2.0 (1.2, 2.9) 37/2,940 | 0.6 (0.3, 0.8) 19/4,202 | 1.0 (0.6, 1.5) 28/4,488 | 0.7 (0.2, 1.2) 17/3,964 |
| Nitrites/poppers | 0.9 (0.5, 1.2) 27/3,192 | 0.8 (0.3, 1.2) 18/3,058 | 0.4 (0.1, 0.7) 10/2,940 | 0.6 (0.3, 0.8) 17/4,202 | 0.1 (0.0, 0.3) 4/4,488 | 0.2 (0.0, 0.5) 3/3,964 |
| Mephedrone | .. | 0.5 (0.1, 0.9) 12/3,058 | 0.1 (0.0, 0.3) 2/2,940 | .. | 0.3 (0.1, 0.5) 8/4,488 | 0.0 0/3,964 |
| Methamphetamine | .. | .. | 0.0 (0.0, 0.1) 1/2,940 | .. | .. | 0.0 0/3,964 |

‘..’ = not asked.

**Table S6 Trends in types of drugs used in the past year by age group and sexual identity: 2007, 2014, 2024**

| | **Heterosexual** | | | **Sexual minority** | | |
|---|---|---|---|---|---|---|
| **Drug type** | **2007** % (95% CI) n/N | **2014** % (95% CI) n/N | **2024** % (95% CI) n/N | **2007** % (95% CI) n/N | **2014** % (95% CI) n/N | **2024** % (95% CI) n/N |
| **Cannabis** | | | | | | |
| 16–24 | 21.3 (17.3, 25.3) 105/498 | 18.4 (14.6, 22.2) 91/509 | 14.8 (9.2, 20.4) 33/239 | 22.4 (10.8, 34.0) 15/63 | 34.9 (18.3, 51.6) 14/38* | 34.1 (19.1, 49.2) 13/43* |
| 25–34 | 13.9 (11.1, 16.7) 117/931 | 11.1 (8.4, 13.8) 97/945 | 14.6 (10.9, 18.3) 80/652 | 28.9 (18.3, 39.5) 25/96 | 17.9 (3.2, 32.7) 8/50 | 16.0 (7.0, 25.1) 13/71 |
| 35–44 | 5.3 (4.0, 6.6) 71/1,296 | 6.4 (4.8, 8.0) 71/1,080 | 9.9 (7.5, 12.3) 84/860 | 16.3 (7.5, 25.0) 17/104 | 26.4 (12.2, 40.7) 12/46* | 10.5 (1.6, 19.3) 7/59 |
| 45–54 | 2.6 (1.7, 3.6) 32/1,037 | 3.5 (2.4, 4.6) 46/1,188 | 7.8 (5.6, 10.0) 73/838 | 8.1 (0.9, 15.3) 6/86 | 7.7 (0.6, 14.8) 5/47* | 23.7 (8.2, 39.2) 9/42* |
| 55–64 | 1.2 (0.5, 1.9) 16/1,209 | 2.1 (1.1, 3.0) 21/1,112 | 3.6 (2.4, 4.7) 44/1,038 | 6.3 (0.0, 13.5) 3/62 | 10.6 (0.0, 21.8) 4/41* | 0.0 0/31* |
| 65+ | 0.3 (0.0, 0.6) 5/1,831 | .. | 1.0 (0.5, 1.5) 24/1,981 | 2.2 (0.0, 5.4) 2/91 | .. | 6.8 (0.0, 17.4) 2/32* |
| **Cocaine** | | | | | | |
| 16–24 | 6.5 (4.4, 8.6) 34/498 | 4.7 (2.7, 6.7) 26/509 | 3.3 (0.4, 6.2) 7/239 | 8.9 (1.5, 16.4) 6/63 | 10.2 (0.0, 23.2) 3/38* | 6.4 (0.0, 15.3) 2/43* |
| 25–34 | 6.0 (4.4, 7.5) 49/931 | 3.7 (2.2, 5.3) 29/945 | 5.0 (2.7, 7.2) 29/652 | 10.8 (2.6, 19.0) 10/96 | 7.9 (0.0, 15.9) 4/50 | 6.8 (0.7, 13.0) 5/71 |
| 35–44 | 1.6 (0.8, 2.3) 21/1,296 | 2.0 (1.1, 2.8) 22/1,080 | 3.5 (1.9, 5.1) 29/860 | 7.8 (0.2, 15.4) 5/104 | 3.1 (0.0, 7.6) 2/46* | 0.0 0/59 |
| 45–54 | 0.3 (0.1, 0.6) 6/1,037 | 0.7 (0.2, 1.2) 9/1,188 | 1.7 (0.5, 2.9) 18/838 | 3.6 (0.0, 9.7) 2/86 | 6.2 (0.0, 13.2) 3/47* | 12.4 (0.0, 25.0) 4/42* |
| 55–64 | 0.0 (0.0, 0.1) 1/1,209 | 0.1 (0.0, 0.2) 1/1,112 | 0.3 (0.0, 0.8) 2/1,038 | 0.0 0/62 | 0.0 0/41* | 0.0 0/31* |
| 65+ | 0.0 0/1,831 | .. | 0.0 (0.0, 0.0) 1/1,981 | 0.0 0/91 | .. | 0.0 0/32* |
| **Ecstasy/MDMA** | | | | | | |
| 16–24 | 3.3 (1.9, 4.8) 19/498 | 6.1 (3.6, 8.5) 28/509 | 3.0 (0.1, 5.9) 6/239 | 4.2 (0.0, 8.5) 4/63 | 6.4 (0.0, 18.6) 1/38* | 10.5 (0.0, 22.2) 3/43* |
| 25–34 | 3.0 (1.8, 4.1) 24/931 | 1.6 (0.6, 2.6) 14/945 | 2.5 (0.7, 4.3) 13/652 | 4.6 (0.1, 9.0) 5/96 | 6.2 (0.0, 13.4) 3/50 | 2.1 (0.0, 4.7) 3/71 |
| 35–44 | 0.6 (0.2, 0.9) 10/1,296 | 1.4 (0.7, 2.1) 15/1,080 | 1.1 (0.3, 1.9) 10/860 | 6.8 (0.0, 13.9) 5/104 | 5.0 (0.0, 11.0) 3/46* | 0.6 (0.0, 1.9) 1/59 |
| 45–54 | 0.0 0/1,037 | 0.1 (0.0, 0.2) 1/1,188 | 0.9 (0.0, 2.0) 6/838 | 0.0 0/86 | 4.3 (0.0, 10.3) 2/47* | 8.7 (0.0, 17.5) 4/42* |
| 55–64 | 0.0 0/1,209 | 0.0 0/1,112 | 0.2 (0.0, 0.5) 2/1,038 | 0.0 0/62 | 0.0 0/41* | 5.9 (0.0, 17.5) 1/31* |
| 65+ | 0.0 0/1,831 | .. | 0.0 (0.0, 0.1) 1/1,981 | 0.0 0/91 | .. | 0.0 0/32* |
| **Ketamine** | | | | | | |
| 16–24 | .. | 2.1 (0.5, 3.7) 9/509 | 3.6 (0.6, 6.6) 7/239 | .. | 6.4 (0.0, 18.6) 1/38* | 8.6 (0.0, 18.4) 3/43* |

| | Heterosexual | | | Sexual minority | | |
|---|---|---|---|---|---|---|
| **Drug type** | **2007** % (95% CI) n/N | **2014** % (95% CI) n/N | **2024** % (95% CI) n/N | **2007** % (95% CI) n/N | **2014** % (95% CI) n/N | **2024** % (95% CI) n/N |
| 25–34 | .. | 0.5 (0.0, 1.2) 3/945 | 1.8 (0.6, 3.1) 10/652 | .. | 1.6 (0.0, 4.9) 1/50 | 0.7 (0.0, 2.2) 1/71 |
| 35–44 | .. | 0.2 (0.0, 0.5) 2/1,080 | 0.6 (0.0, 1.3) 4/860 | .. | 0.0 0/46* | 1.1 (0.0, 2.6) 2/59 |
| 45–54 | .. | 0.0 0/1,188 | 0.0 (0.0, 0.1) 1/838 | .. | 0.0 0/48* | 0.0 0/42* |
| 55–64 | .. | 0.0 0/1,112 | 0.0 0/1,038 | .. | 0.0 0/41* | 0.0 0/31* |
| 65+ | .. | .. | 0.0 0/1,981 | .. | .. | 0.0 0/32* |
| **Nitrites/poppers** | | | | | | |
| 16–24 | 1.8 (0.6, 3.0) 9/498 | 1.3 (0.1, 2.5) 5/509 | 0.0 0/239 | 5.9 (0.6, 11.3) 5/63 | 6.4 (0.0, 18.6) 1/38* | 7.0 (0.0, 15.5) 3/43* |
| 25–34 | 1.0 (0.3, 1.6) 10/931 | 0.5 (0.0, 1.0) 3/945 | 0.1 (0.0, 0.4) 1/652 | 1.3 (0.0, 3.1) 2/96 | 3.2 (0.0, 7.8) 2/50 | 0.0 0/71 |
| 35–44 | 0.7 (0.2, 1.1) 10/1,296 | 0.1 (0.0, 0.2) 1/1,080 | 0.2 (0.0, 0.5) 2/860 | 3.1 (0.0, 6.2) 4/104 | 10.8 (0.1, 21.6) 4/46* | 0.5 (0.0, 1.4) 1/59 |
| 45–54 | 0.0 0/1,037 | 0.1 (0.0, 0.3) 1/1,188 | 0.2 (0.0, 0.5) 1/838 | 4.4 (0.0, 10.9) 2/86 | 4.2 (0.0, 9.5) 3/48* | 6.0 (0.0, 15.3) 2/42* |
| 55–64 | 0.0 0/1,209 | 0.0 0/1,112 | 0.1 (0.0, 0.2) 1/1,038 | 1.8 (0.0, 5.3) 1/62 | 3.8 (0.0, 9.2) 2/41* | 0.0 0/31* |
| 65+ | 0.0 0/1,831 | .. | 0.0 0/1,981 | 0.8 (0.0, 2.4) 1/91 | .. | 4.4 (0.0, 13.1) 1/32* |
| **Mephedrone** | | | | | | |
| 16–24 | .. | 0.8 (0.1, 1.6) 6/509 | 0.0 0/239 | .. | 8.7 (0.0, 21.6) 2/38* | 0.0 0/43* |
| 25–34 | .. | 0.6 (0.1, 1.2) 5/945 | 0.0 0/652 | .. | 3.1 (0.0, 7.5) 2/50 | 2.0 (0.0, 5.9) 1/71 |
| 35–44 | .. | 0.0 (0.0, 0.1) 1/1,080 | 0.2 (0.0, 0.7) 1/860 | .. | 2.4 (0.0, 7.2) 1/46* | 0.0 0/59 |
| 45–54 | .. | 0.0 0/1,188 | 0.0 0/838 | .. | 5.7 (0.0, 13.9) 2/48* | 0.0 0/42* |
| 55–64 | .. | 0.0 0/1,112 | 0.0 0/1,038 | .. | 0.0 0/41* | 0.0 0/31* |
| 65+ | .. | .. | 0.0 0/1,981 | .. | .. | 0.0 0/32* |
| **Methamphetamine** | | | | | | |
| 16–24 | .. | .. | 0.0 0/239 | .. | .. | 0.0 0/43* |
| 25–34 | .. | .. | 0.0 0/652 | .. | .. | 2.0 (0.0, 5.9) 1/71 |
| 35–44 | .. | .. | 0.0 0/860 | .. | .. | 0.0 0/59 |

| | Heterosexual | | | Sexual minority | | |
|---|---|---|---|---|---|---|
| **Drug type** | **2007** % (95% CI) n/N | **2014** % (95% CI) n/N | **2024** % (95% CI) n/N | **2007** % (95% CI) n/N | **2014** % (95% CI) n/N | **2024** % (95% CI) n/N |
| 45–54 | .. | .. | 0.0 0/838 | .. | .. | 0.0 0/42* |
| 55–64 | .. | .. | 0.0 0/1,038 | .. | .. | 0.0 0/31* |
| 65+ | .. | .. | 0.0 0/1,981 | .. | .. | 0.0 0/32* |

'*' Note small base size for these estimates (unweighted N <50).
'..' = not asked.

### Figures

**Fig. S1 Prevalence of past-year drug use by sexual identity among adults aged 16+, APMS 2007, 2024**

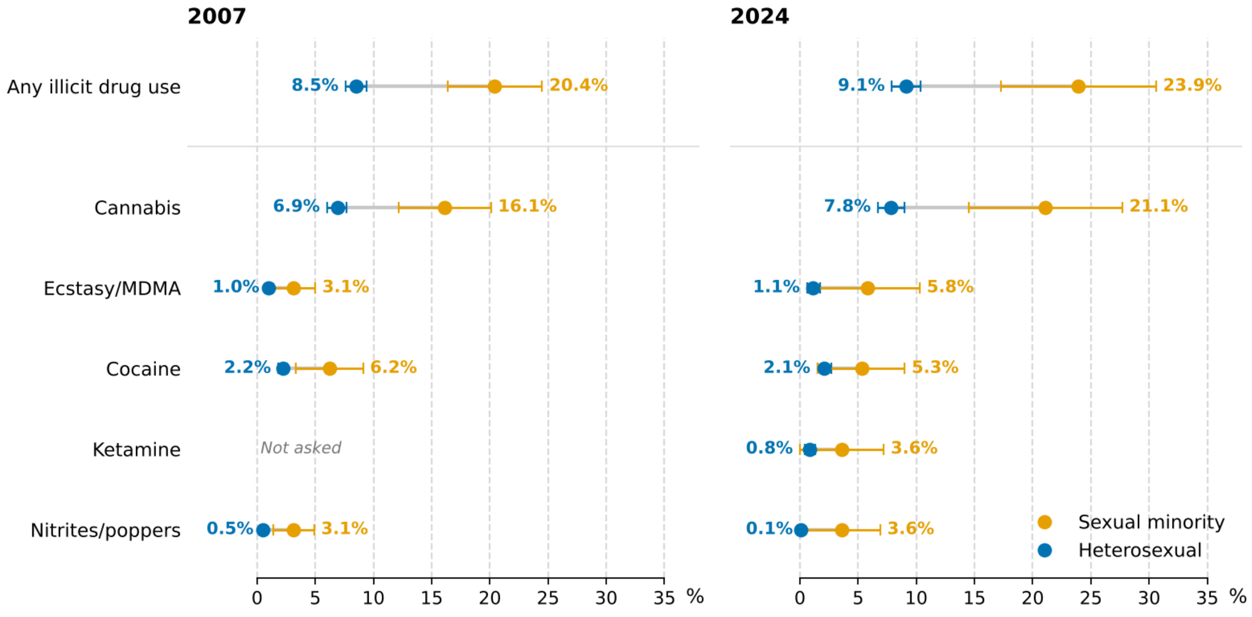
